# Proposed Cuts to the Ryan White Program Could Raise HIV Incidence by 18% in 30 US States and the District of Columbia: A Simulation Study

**DOI:** 10.1101/2025.10.14.25337745

**Authors:** Andrew Zalesak, Melissa Schnure, Ryan Forster, Joyce L. Jones, Catherine R. Lesko, D. Scott Batey, Keri N. Althoff, Kelly A. Gebo, David W. Dowdy, Maunank Shah, Parastu Kasaie, Anthony T. Fojo

## Abstract

Potential cuts to Ryan White Parts C and D, and funding for the Minority AIDS and EHE threaten to undermine the progress made in controlling the US HIV epidemic. Using an HIV simulation model in 30 states and Washington, DC, we project that ending these programs would lead to 23,883 additional HIV infections over five years - a 17.6% increase compared to continuing all Ryan White programs. The projected increases varied by state but were disproportionately large in the seven EHE priority states with a high rural burden of HIV. These findings highlight the importance of Ryan White services in preventing HIV transmission in the US.

## Background

The federal Ryan White HIV/AIDS Program provides HIV treatment and support services for half of the 1.2 million people with HIV in the United States, many of whom would not otherwise be able to access care^1^. Part A provides grants to city and county health departments. Part B provides grants to states and includes the AIDS Drug Assistance Program (ADAP) which pays for antiretroviral medications directly or indirectly (by paying for insurance or medication copays). Part C supports community groups to provide outpatient ambulatory health services and support. Part D funds outpatient services specific for women, infants, and youth. Part F supports dental care and professional education. The Minority AIDS Initiative works to improve outcomes for racial and ethnic minority populations disproportionately affected by HIV. Since 2019, the US Ending the HIV Epidemic (EHE) initiative provides Ryan White funds to seven priority states with a high rural burden of HIV, and 50 high burden urban counties^2^.

Except for ADAP and Part F, most Ryan White programs facilitate outpatient medical care and provide supportive services such as case management, transportation, and housing assistance. These programs are instrumental in helping people with HIV maintain viral suppression; when virally suppressed, they cannot transmit HIV^2^.

### Objective

We sought to estimate the excess HIV infections that would result from potential cuts to Ryan White Parts C and D, and allocations to the Minority AIDS and EHE initiatives in 30 US states and Washington, DC.

## Methods and Findings

We used the Johns Hopkins Epidemiologic and Economic Model, a dynamic HIV transmission model (Supplement Figure 1), which we have previously used to project the impacts of potential cuts to the entire Ryan White program^3^. In this analysis, we categorized clients into two groups: (1) those receiving outpatient ambulatory health services, with or without other services, and (2) those receiving only support services without ambulatory health services.

For each location, we calculated the state-level fraction of all non-ADAP Ryan White funds that came from Parts C, D, and the Minority AIDS and EHE initiatives (Supplement Table 1). We presumed that this would approximate the fraction of Ryan White clients who would lose outpatient health and support services from cuts to these programs. For those who lose services, we simulated the proportion who would lose viral suppression based on a survey of Ryan White clinic directors and administrators, as in our previous studies^3^, varied across 1,000 simulations in each state.

We simulated two scenarios where programs supported by Parts C, D, and the Minority Health and EHE initiatives end in February 2026. In our “Cessation” scenario, services never return; in our “Interruption” scenario, services return in January 2029, and viral suppression among Ryan White clients recovers by December 2029.

If Ryan White services continue, we project 135,745 new HIV infections (95% credible interval: 130,450 to 141,621) from 2026-2030 across all 30 states and DC. If services supported by Parts C, D, and Minority Health and EHE funds end permanently in February 2026, we project 23,883 additional infections (2,812 to 53,813) from 2026-2030, an excess of 17.6% (2.0 to 39.5%). If programs are restored in January 2029, we project 17,710 additional infections (2,073 to 39,829) from 2026-2030, an excess of 13.1% (1.5 to 29.3%). The expected increases in HIV infections vary across states (see Figure), ranging from 4.4% (0.3 to 11.0%) in Indiana to 53.8% (8.9 to 112.9%) in South Carolina. Four of the five states with the highest increases infections were EHE priority states, and the other three EHE priority states were in the top half.

## Discussion

Using an HIV simulation model in 30 states and Washington, DC, we project that ending Ryan White Parts C and D, and funding for the Minority AIDS and EHE initiatives would lead to 23,883 additional HIV infections over five years – a 17.6% increase compared to continuing all Ryan White programs. The projected increases varied by state but were disproportionately large in the seven EHE priority states with a high rural burden of HIV.

These findings highlight the importance of Ryan White services in preventing HIV transmission in the US. The projected rise in infections resulting from service cuts threatens to undermine the progress made in controlling the US HIV epidemic.

**Figure:**
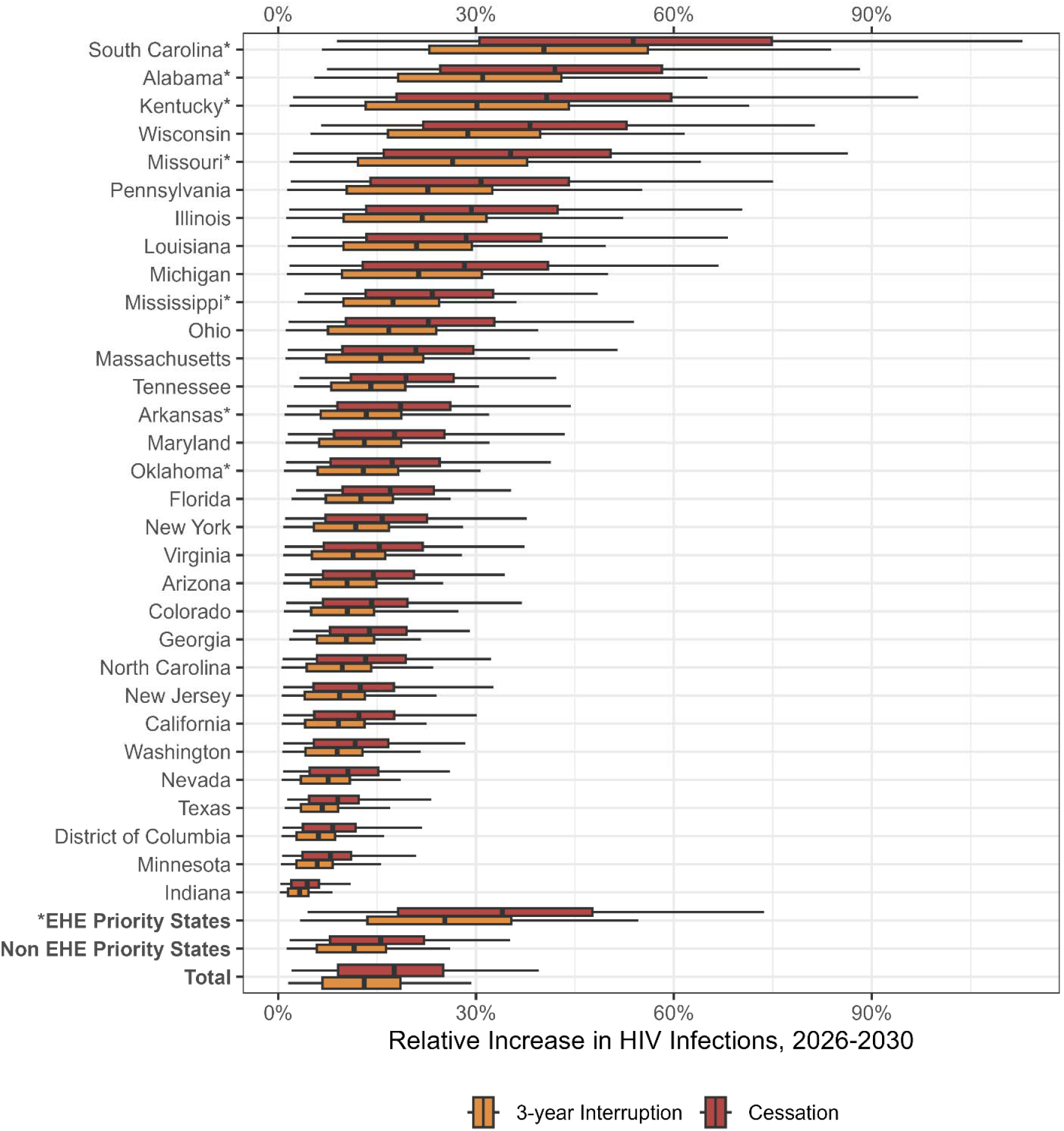
Relative Excess HIV Infections from 2025-2030 If Ryan White HIV/AIDS Program Part C, D, Minority AIDS Initiative, and Ending the HIV Epidemic Initiative Programs are Stopped or Interrupted in 30 States and Washington, DC. Boxplots display the percentage increase in new HIV infections from 2026 to 2030 under two scenarios in which certain Ryan White services (Parts C, D, Minority AIDS Initiative and Ending the HIV Epidemic Initiative funding) stop on February 1, 2026, relative to continuing Ryan White services. Red denotes “Cessation,” in which viral suppression among Ryan White clients never recovers; orange denotes “Interruption,” in which viral suppression among Ryan White clients recovers from January to December 2029. The value along the x-axis represents the relative increase in cases vs. a scenario where Ryan White services continue uninterrupted. The dark vertical lines indicate the median projection across 1,000 simulations, the boxes indicate interquartile ranges (IQR), and whiskers cover the 95% credible interval. EHE priority states are labeled with an asterisk.

## Supporting information

Technical Supplement

## Data Availability

All data produced in the present study are available upon reasonable request to the authors.

## Notes

### Competing Interest Statement

The authors have declared no competing interest.

### Funding Statement

This research was supported by NIH Grant Number R01MD018539.

