## Supplementary material for "Proposed Cuts to the Ryan White Program Could Raise HIV Incidence by 18% in 30 US States and the District of Columbia: A Simulation Study": Technical Supplement

1. Andrew Zalesak, MS

Johns Hopkins University School of Medicine

2. Melissa Schnure, PhD ScM

Johns Hopkins University School of Medicine

3. Ryan Forster, PhD

Johns Hopkins Bloomberg School of Public Health

4. Joyce L. Jones, MD MS

Johns Hopkins University School of Medicine

5. Catherine R. Lesko, PhD

Johns Hopkins Bloomberg School of Public Health

6. D. Scott Batey, PhD MSW

Tulane University School of Social Work

7. Keri N. Althoff, PhD MPH

Johns Hopkins Bloomberg School of Public Health

8. Kelly A. Gebo, MD MPH

Johns Hopkins University School of Medicine

9. David W. Dowdy, MD PhD

Johns Hopkins Bloomberg School of Public Health

10. Maunank Shah, MD PhD

Johns Hopkins University School of Medicine

11. Parastu Kasaie, PhD MS

Johns Hopkins Bloomberg School of Public Health

12. Anthony T. Fojo, MD MHS

Johns Hopkins University School of Medicine

Model Structure

**Figure S1: Model Structure**

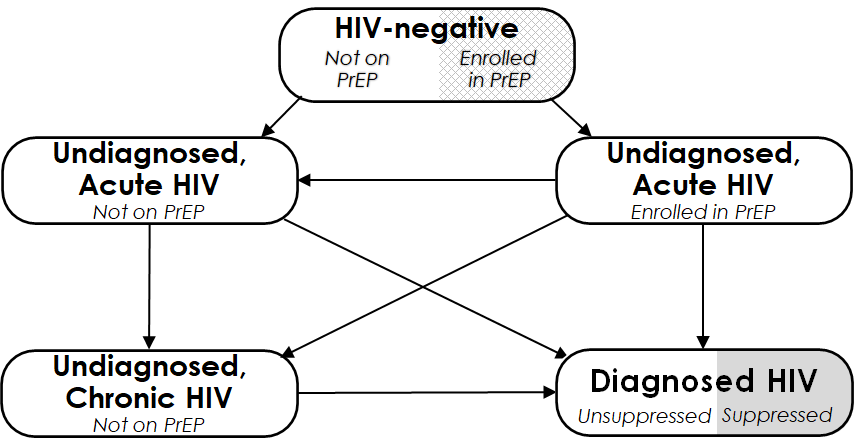

This figure depicts the compartments representing HIV status. Each of the five compartments is further stratified by age (13–24, 25–34, 35–44, 45–54, and ≥55 years), race/ethnicity (Black, Hispanic, and other), sex and sexual behavior (female, heterosexual male, and men who have sex with men (MSM), and intravenous drug use history (never used, active use, and prior use). “Acute HIV” refers to the first 2.9 months following infection, during which risk of transmission is high.

Ryan White Costs

**Table S1: Ryan White FY 2023 Cost Data (USD)**

| **State** | **Part A** | **Part B non-ADAP** | **Part C** | **Part D** | **EHE** | **Minority AIDS Initiative from parts A and B** | **Total non-ADAP** | **Total Potential Funding Cut** | **Percent Potential Funding Cut (%)** |
| --- | --- | --- | --- | --- | --- | --- | --- | --- | --- |
| Alabama | - | 9,403,840 | 3,576,316 | 1,923,127 | 2,555,761 | 157,830 | 17,459,044 | 8,213,034 | 47 |
| Arizona | 10,747,912 | 4,881,358 | 1,940,930 | 680,985 | 2,555,761 | 797,250 | 20,806,946 | 5,974,926 | 29 |
| Arkansas | - | 3,829,697 | 1,113,916 | 924,967 | 2,000,000 | 53,985 | 7,868,580 | 4,092,868 | 52 |
| California | 104,115,890 | 49,980,378 | 20,047,529 | 5,768,033 | 21,548,760 | 8,704,751 | 201,460,590 | 56,069,073 | 28 |
| Colorado | 7,798,172 | 7,379,471 | 1,735,138 | 878,135 | - | 477,810 | 17,790,916 | 3,091,083 | 17 |
| District of Columbia | 32,652,189 | 3,683,371 | 2,248,879 | - | 3,755,939 | 3,001,061 | 42,340,378 | 9,005,879 | 21 |
| Florida | 79,821,788 | 46,091,194 | 11,415,428 | 7,182,408 | 16,721,032 | 7,883,218 | 161,231,850 | 43,202,086 | 27 |
| Georgia | 31,327,260 | 28,974,070 | 11,480,290 | 1,962,913 | 5,079,513 | 3,418,037 | 78,824,046 | 21,940,753 | 28 |
| Illinois | 27,842,975 | 10,405,191 | 6,955,171 | 2,848,918 | 4,647,156 | 2,754,620 | 52,699,411 | 17,205,865 | 33 |
| Indiana | 4,841,732 | 15,730,885 | 1,300,314 | - | 2,000,000 | 326,526 | 23,872,931 | 3,626,840 | 15 |
| Kentucky | - | 5,138,823 | 2,929,299 | 1,380,108 | 2,000,000 | 50,992 | 11,448,230 | 6,360,399 | 56 |
| Louisiana | 12,925,451 | 7,420,281 | 5,614,251 | 2,878,348 | 4,000,000 | 1,321,764 | 32,838,331 | 13,814,363 | 42 |
| Maryland | 16,350,167 | 10,346,982 | 2,541,368 | 1,354,120 | 2,878,672 | 1,865,530 | 33,471,309 | 8,639,690 | 26 |
| Massachusetts | 15,228,608 | 7,011,076 | 6,640,998 | 1,909,518 | 2,854,962 | 1,198,414 | 33,645,162 | 12,603,892 | 37 |
| Michigan | 10,135,391 | 6,299,164 | 2,818,189 | 1,806,383 | 2,406,807 | 1,004,741 | 23,465,934 | 8,036,120 | 34 |
| Minnesota | 6,193,009 | 5,241,046 | 445,228 | 562,092 | - | 389,269 | 12,441,375 | 1,396,589 | 11 |
| Mississippi | - | 6,659,820 | 2,972,766 | 587,231 | 2,195,932 | 127,383 | 12,415,749 | 5,883,312 | 47 |
| Missouri | 11,011,849 | 3,829,578 | 2,466,217 | 1,948,823 | 2,555,761 | 760,161 | 21,812,228 | 7,730,962 | 35 |
| Nevada | 7,225,692 | 5,984,615 | 1,782,051 | 607,114 | 2,015,473 | 507,424 | 17,614,945 | 4,912,062 | 28 |
| New Jersey | 24,436,681 | 16,091,669 | 5,207,780 | 2,150,640 | 4,555,761 | 2,615,401 | 52,442,531 | 14,529,582 | 28 |
| New York | 99,065,139 | 44,031,208 | 20,521,029 | 8,082,708 | 16,750,409 | 9,950,107 | 188,450,493 | 55,304,253 | 29 |
| North Carolina | 6,714,279 | 24,926,783 | 7,221,281 | 3,848,811 | - | 964,274 | 42,711,154 | 12,034,366 | 28 |
| Ohio | 9,873,562 | 8,324,755 | 3,915,785 | 896,185 | 6,000,000 | 685,166 | 29,010,287 | 11,497,136 | 40 |
| Oklahoma | - | 4,390,387 | 1,676,908 | 534,587 | 2,000,000 | - | 8,601,882 | 4,211,495 | 49 |
| Pennsylvania | 23,171,888 | 11,809,043 | 11,516,779 | 3,235,879 | 3,156,217 | 2,263,628 | 52,889,806 | 20,172,503 | 38 |
| South Carolina | - | 11,801,358 | 5,823,100 | 1,196,995 | 4,810,331 | 209,879 | 23,631,784 | 12,040,305 | 51 |
| Tennessee | 11,655,851 | 16,166,802 | 2,824,359 | 1,152,061 | 2,000,000 | 1,185,352 | 33,799,073 | 7,161,772 | 21 |
| Texas | 64,476,652 | 40,974,275 | 10,093,899 | 5,623,717 | 14,054,421 | 6,478,921 | 135,222,964 | 36,250,958 | 27 |
| Virginia | 6,020,029 | 8,329,308 | 2,794,884 | 1,060,867 | - | 793,256 | 18,205,088 | 4,649,007 | 26 |
| Washington | 7,362,672 | 4,124,910 | 1,122,124 | 434,758 | 2,000,000 | 472,687 | 15,044,464 | 4,029,569 | 27 |
| Wisconsin | - | 4,272,795 | 2,230,338 | 922,890 | - | 56,173 | 7,426,023 | 3,209,401 | 43 |

Ryan White grant awards in fiscal year 2023 by state. “Part B non-ADAP” funding was calculated by subtracting the AIDS Drug Assistance Program (ADAP) award value from the total Part B award value. “Total non-ADAP” funding was calculated by subtracting the Part B ADAP award from the total Ryan White award. “Total Potential Cut” is the sum of Part C, Part D, EHE, and Minority AIDS Initiative funding from parts A and B. “Percent Potential Funding Cut” was calculated by dividing the total potential funding cut by the total non-ADAP award. Sources: Part A^1^, Part B^2^, Part C^3,4^, Part D^5^, EHE^6^.
